# Multimodal deep learning enhances diagnostic precision in left ventricular hypertrophy

**DOI:** 10.1101/2021.06.13.21258860

**Authors:** Jessica Torres Soto, J. Weston Hughes, Pablo Amador Sanchez, Marco Perez, David Ouyang, Euan Ashley

## Abstract

Determining the etiology of left ventricular hypertrophy (LVH) can be challenging due to the similarity in clinical presentation and cardiac morphological features of diverse causes of disease. In particular, distinguishing individuals with hypertrophic cardiomyopathy (HCM) from the much larger set of individuals with manifest or occult hypertension (HTN) is of major importance for family screening and the prevention of sudden death. We hypothesized that deep learning based joint interpretation of 12 lead electrocardiograms and echocardiogram videos could augment physician interpretation. We chose not to train on proximate data labels such as physician over-reads of ECGs or echocardiograms but instead took advantage of electronic health record derived clinical blood pressure measurements and diagnostic consensus (often including molecular testing) among physicians in an HCM center of excellence. Using over 18,000 combined instances of electrocardiograms and echocardiograms from 2,728 patients, we developed LVH-Fusion. On held-out test data, LVH-Fusion achieved an F1-score of 0.71 in predicting HCM, and 0.96 in predicting HTN. In head-to-head comparison with human readers LVH-Fusion had higher sensitivity and specificity rates than its human counterparts. Finally, we use explainability techniques to investigate local and global features that positively and negatively impact LVH-Fusion prediction estimates providing confirmation from unsupervised analysis the diagnostic power of lateral T wave inversion on the ECG and proximal septal hypertrophy on the echocardiogram for HCM. In conclusion, these results show that deep learning can provide effective physician augmentation in the face of a common diagnostic dilemma with far reaching implications for the prevention of sudden cardiac death.

## Introduction

Hypertrophic cardiomyopathy (HCM) is the most common cardiac genetic disease with an estimated prevalence in the general population of 1:500 to 1:200.^1^ HCM is an autosomal dominant mendelian disease that can be associated with significant morbidity in the form of heart failure and sudden death.^2^ Thus, identifying patients with HCM has significance well beyond the individual, with many proband diagnoses leading to screening of several generations of a family. Diagnosis of HCM can be difficult due to the high prevalence of manifest hypertension in the general population, present in up to 45% of US adults ^3^ (this before counting the occult disease). Thus, a common diagnostic dilemma for clinicians when faced with LVH on the ECG or echocardiogram is how to rule out HCM. In a small study, the rates of misclassification of HCM were as high as 30% percent with hypertension being the most common misdiagnosis ^4^. Although the American Heart Association provides guidelines for the diagnosis of hypertension and HCM separately, distinguishing between them is a task that most physicians feel ill equipped to perform (understandably as HCM is a rare disease not commonly encountered even in general cardiology practice). This provides an opportunity for physician augmentation through artificial intelligence (AI).

New advances in artificial intelligence have led to rapid expansion of medical deep learning applications with an emphasis on medical specialties that hold a high degree of visual pattern recognition tasks like radiology, pathology, ophthalmology, dermatology and most notably cardiology.^5^ Imaging and electrical phenotypes of hypertrophic cardiomyopathy ^6,7^ are the first line clinical tools.

Interpretation of the ECG relies on direct visual assessment making it ideal for deep learning approaches. Previous work has demonstrated that demographic and medical data can be learned including detection of low ejection fraction, something typically requiring echocardiography to confirm ^8–11^. Our prior work using video computation of echocardiograms has demonstrated efficient detection of left ventricular hypertrophy and the identification of a broad range of cardiovascular disease.^12,13^

Combining data sources as human diagnosticians do, has the potential to provide an artificial intelligence (AI) algorithm with greater diagnostic power^14^. We focus here on the two most frequent diagnostic modalities in cardiology. To date, no published work has explored the benefits of a multimodal deep learning model using electrocardiogram and echocardiogram data, although there has been some exploration of combining separately trained diagnostic models in a single pipeline^15^. We hypothesize that multimodal deep learning may provide added benefit in distinguishing patterns that are not easily discernible from individual modalities. We present LVH-fusion, the first model to jointly model electrical and ultrasound-based time series data of the heart. We demonstrate its potential with application to the diagnosis of left ventricular hypertrophy.

## Results

We developed a multi-modal deep learning framework, LVH-fusion, that takes as input time based electrical and echocardiographic data of the heart. We applied this framework in a common clinical challenge: the determination of the etiology of left ventricular hypertrophy. Motivated by prior work on deep learning applied to electrocardiogram signals and echocardiogram videos ^9,13,16^, LVH-fusion jointly models both electrocardiogram and echocardiogram data. It is trained not with proximate human derived ECG and echocardiogram labels but rather via a gold standard diagnosis independently derived from the Electronic Health Records (HTN) or through the consensus diagnosis of HCM within a center of excellence.

In this study, both single-modal and multimodal neural network models were examined (Figure 1). Four different multimodal fusion architectures were explored, combining ECG and echocardiogram information in different ways. For both late-average fusion and late-ranked fusion models, decision level fusion was used to combine the outputs of electrocardiogram and echocardiogram classifiers^17^. In the late-average fusion model, soft voting is performed by computing the average probability for each class from the individual ECG and echocardiogram classifiers and predicts the class with maximal average probability. In the late-ranked fusion model, the probabilities for each class from the individual ECG and echocardiogram classifiers are ranked and a prediction is determined from the highest ranked probability. For the late fusion models, both pre-trained and random, the learned feature representations from each modality were concatenated together before the final classification layer. In this situation the fusion model considers both inputs and during training and the loss is calculated jointly. We explored the benefits of randomly initialized weights and pretrained weights in the late fusion model. Lastly, the single modal models provide a benchmark against which to compare multimodal models that jointly consider the paired electrocardiogram and echocardiogram data, demonstrating the benefit of a combined approach.

**Figure 1.**
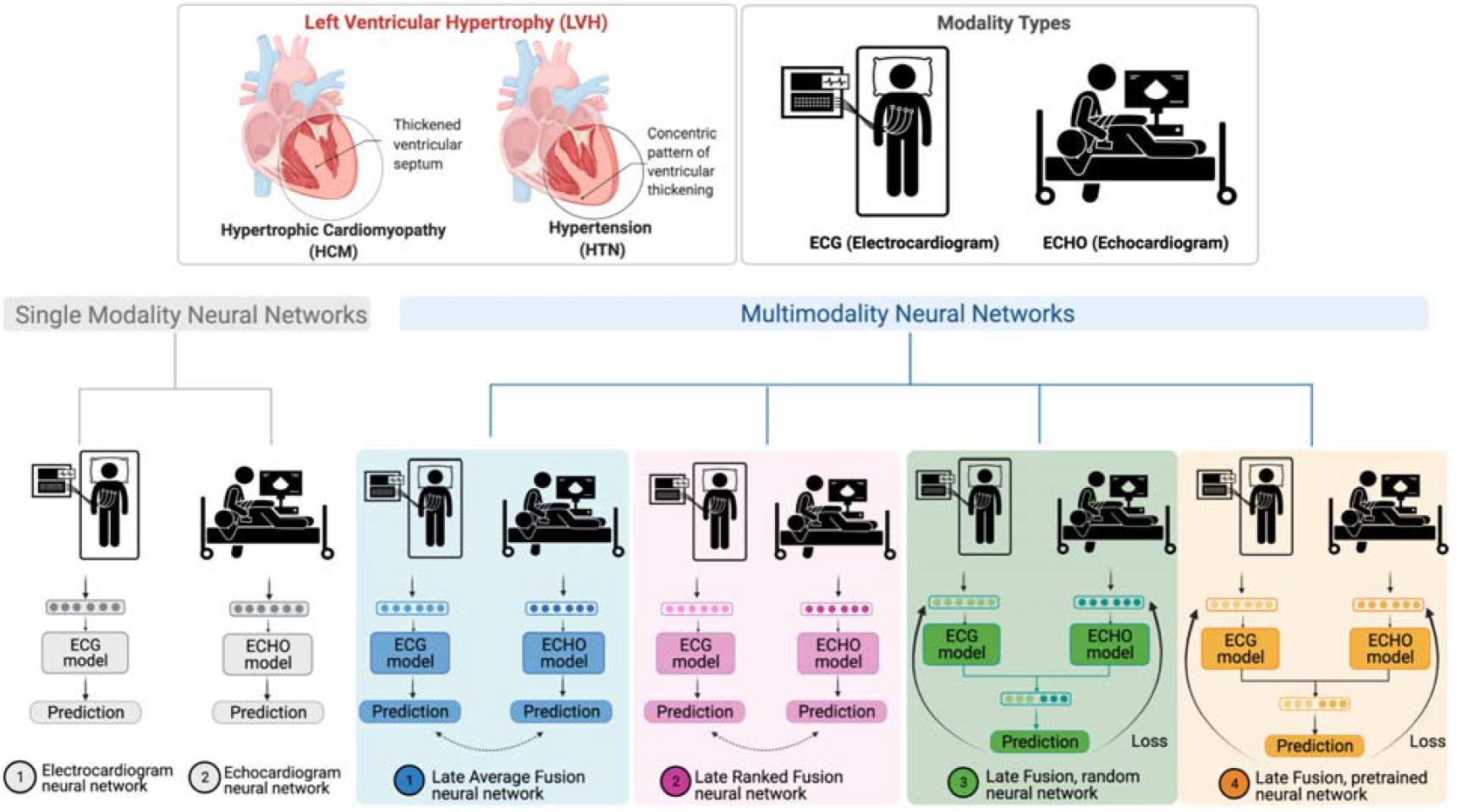
LVH-Fusion study design. Two disease of interested are denoted, HCM and HTN, alongside data modality types used in this study. Single modal as well as multimodal model architecture were explored. LVH-Fusion is based on a late average fusion neural network, denoted in blue.

### Data Acquisition and selection

With the approval of Stanford Institutional Review Board (IRB), we retrieved electrocardiograms and echocardiograms from patients between 2006 and 2018 at Stanford Medicine (Table 1). The data was split into training, validation, and test sets with no patient overlap between sets. Due to the fact that multiple electrocardiograms and echocardiograms are present within the healthcare system record, we explored various data selection scenarios to understand what selection methods are best suited for this specific task. The quantitative comparison of all data selection used can be found in Supplementary Table S1. The final model was trained using a patient’s first ECG and first echocardiogram in the system.

**Table 1.** Breakdown of data by partition.

| Label | Partition | Number of unique<br>Echo patients | Number of<br>Echos | Number of unique<br>ECG patients | Number of<br>ECGs |
| --- | --- | --- | --- | --- | --- |
| HCM | Train | 256 | 596 | 662 | 4,281 |
|  | Validate | 31 | 58 | 71 | 424 |
|  | Test | 27 | 88 | 78 | 380 |
| HTN | Train | 1,469 | 1,976 | 1,535 | 8,348 |
|  | Validate | 186 | 270 | 191 | 1,127 |
|  | Test | 181 | 246 | 191 | 1,201 |

### Model performance

Four multimodal fusion models were explored: late-average, late-ranked, pre-trained late fusion and random late fusion (Figure 1). The performance metrics of each model is detailed in Table 2. The late average model achieved the highest F1-score and specificity rates 0.711 (0.571 - 0.826) and 0.952 (0.921 - 0.979) respectively on the held-out test set. We conducted experiments to study the performance of single-modal models trained on only ECG and echo to demonstrate the benefit of multimodal models. The multimodal models outperform single-modal model F1-scores, which increase from 0.63 to 0.71. Furthermore, the false-discovery rates are significantly reduced from 0.45 to 0.3. To provide context for these results, we also trained the single-modal models to predict left ventricular etiology using standard quantitative features from the electrocardiogram. This baseline model achieved sensitivity rates of 0.51 for predicting HCM which is considerably lower than LVH-Fusion (Supplementary Table S2). These results show that the proposed electrocardiogram signals model discover novel characteristics not accounted for with the quantitative features. Lastly, to examine the discriminatory power of our methodology, we performed a sensitivity analysis for predicting LVH etiology including the additional classification task of “normal.” In this context, LVH-fusion maintains high discriminatory power in predicting LVH from normal ECG and echocardiogram videos, suggesting that false positive rates of hypertension or hypertrophic cardiomyopathy would be low if the model was extended to this use case (Supplementary Table S3 and S4).

**Table 2:**
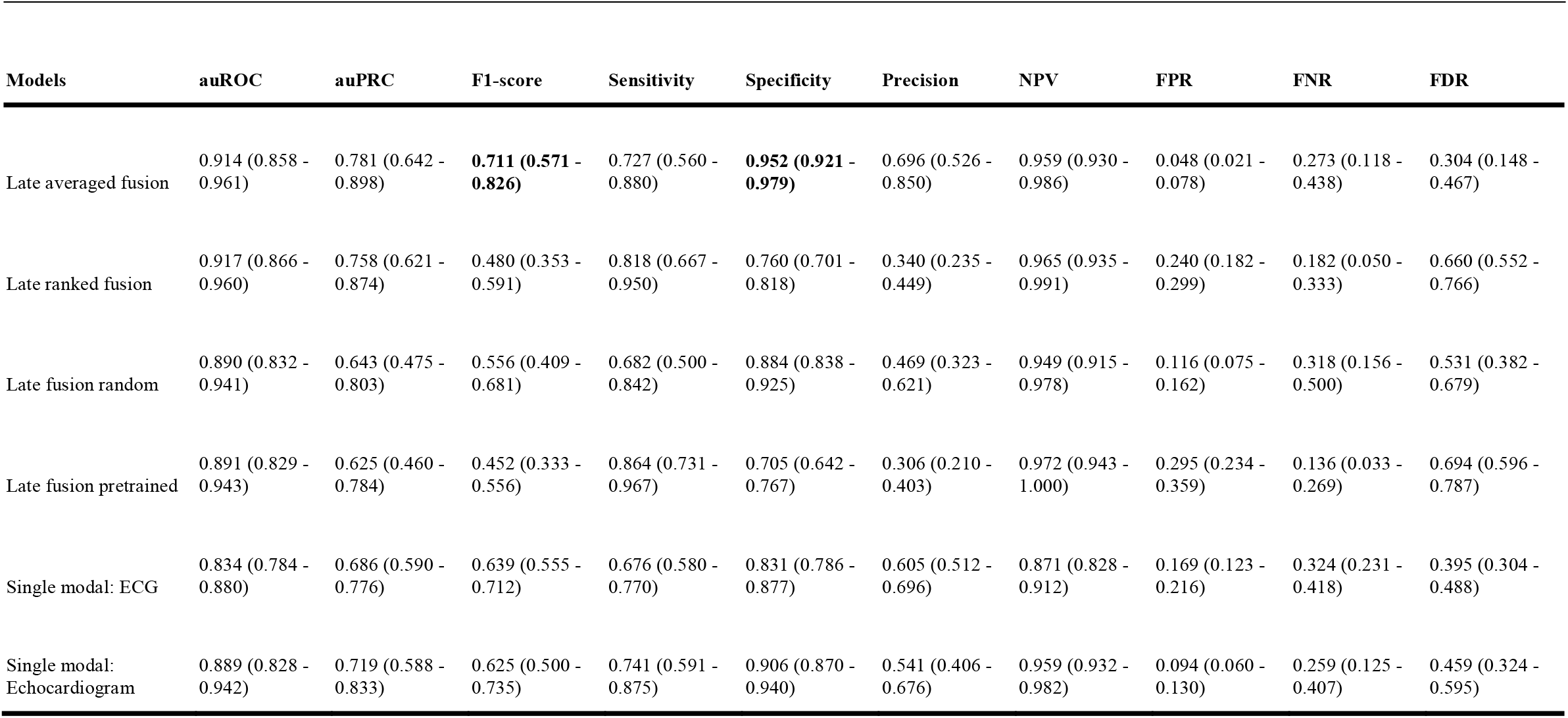
Model performance metrics.

### Understanding model performance

In order to improve our understanding of how LVH-Fusion classifies left ventricular etiology, we implemented a series of ablation studies similar to Hughes et al. 2021^18^ to determine what information models rely on to make predictions. For electrocardiogram single-modal models we examined the impact of varying the number of leads from the standard 12 leads to 8 leads and masking each lead to understand the impact each lead holds for prediction estimates. We find that although no single lead harbors a statistically significant impact on the overall model performance, masking out lead V3 and aVR had the highest negative impact on prediction estimates, Figure 2. Next, since the standard 12 lead ECG contains 8 algebraically independent leads, we considered the impact of masking multiple leads combinations. We observe an overall reduction in classification metrics when masking multiple leads at a time with no significant difference between masking the 4 dependent leads (III, aVL, aVF, aVR) and a random subselection of 4 leads, Supplementary Figure S2. These results suggest our model benefits from the complete 12 lead input and classification metrics are negatively impacted with any nonspecific reduction in leads.

**Figure 2.**
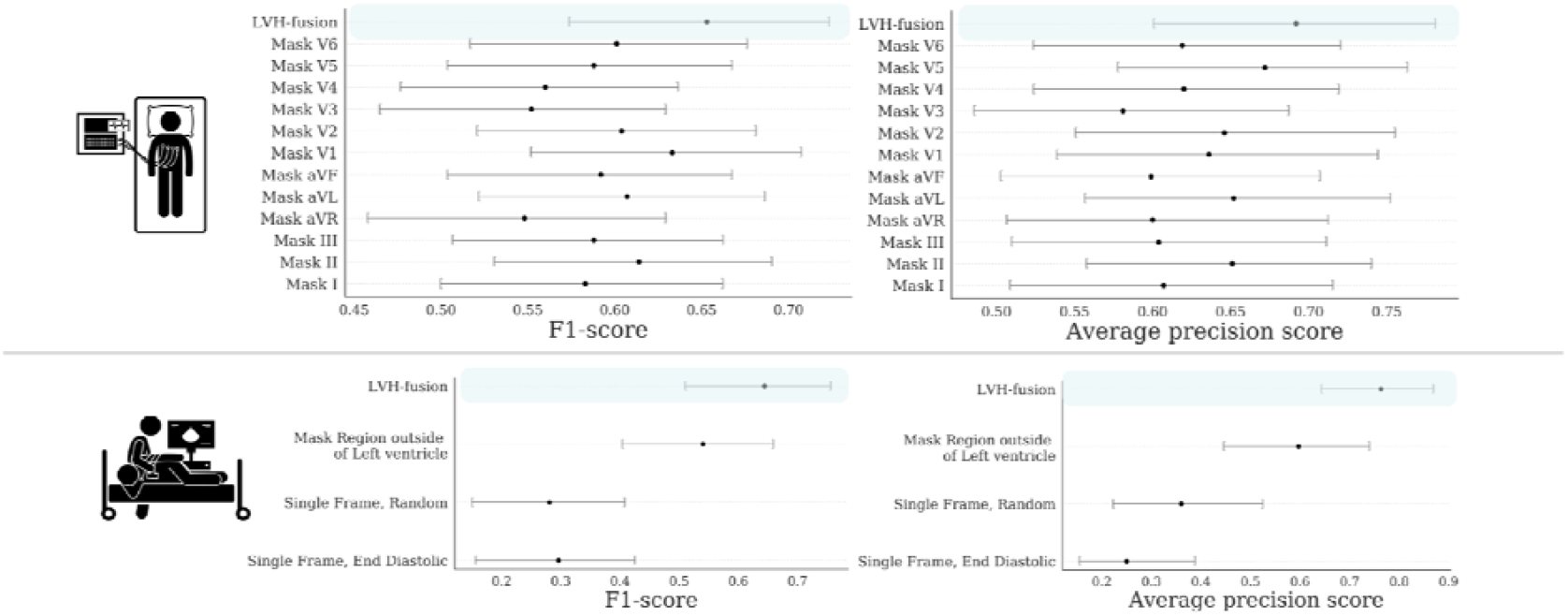
Ablation studies impact on LVH-Fusion performance. Bootstrap 95% CI for performance metrics, F1-score and average precision score, for each model trained on ablated input data. for each prediction metric is shown. (TOP row) Results from ablating ECG input. (BOTTOM row) Results from ablating echocardiogram input. For each ablation setting, a separate model was trained on that type of ablated data to quantify the information content in the data.

For the echocardiogram single-modal model, we examined segmentation, restricting the prediction algorithm to i) only the region around the left ventricle, ii) random single frames, and iii) single end diastolic frames. Restricting the echocardiogram model to the area around the left ventricle caused a decrease in accuracy, showing the model relies on information outside of that region to make classifications. This is interesting given the focus of clinicians on the left ventricle when considering LVH, even despite the fact that hypertension could impact the left atrium by causing restriction and HCM affects all four chambers. Restricting the model’s input to a single frame further decreases accuracy, demonstrating that motion information is important in distinguishing between HCM and hypertension. Figure 2 details the performance of each ablation experiment.

### Model interpretations

In order to improve our understanding of how LVH-Fusion classifies left ventricular etiology, we implemented SHAP GradientExplainer, a game theory approach to explain the output of a machine learning algorithm^19^. Relating this method to the ECG model, this approach takes the prediction of a model and estimates the gradient with respect to each individual timestep for every lead from the input signal. For echocardiogram videos, an analogous methodology applies: the gradient of the model’s prediction was calculated with respect to every pixel from the input video. In each case, the calculated value is then compared to a provided background distribution, the training data. The value of the calculated gradients for each timestep/pixel is then assigned an importance score such that highly impactful scores (denoted in red) hold positive impacts on prediction estimates. Values with low importance scores negatively influence prediction estimates (denoted in blue).

We emphasize samples of ECG and echocardiograms from the test partition to deduce regions the model found most impactful to prediction estimates, Figure 3 and 4. In Figure 3, the ECG interpretation results highlight an overall focus on V3 and T-wave inversion in leads V1-V6. Both the observed early R wave progression and T-wave inversion are indications of HCM. Summarized local interpretations for each lead provides explanations of the overall impact each lead has on prediction estimates. Additional examples of ECG interpretation tracings can be found in the Supplement Figure S1. Comparably, the interpretation results of the echocardiogram videos, Figure 4, clearly depicts asymmetric proximal septal thickness, a hallmark distinction of HCM across all frames of the video. Next, to examine local summary interpretations, we segmented the left ventricle on each frame for duration of a video’s length. This allowed us to quantitatively compare the positive and negative impacts the estimated LV size had on overall prediction estimates, Supplemental Figure S3.

**Figure 3.**
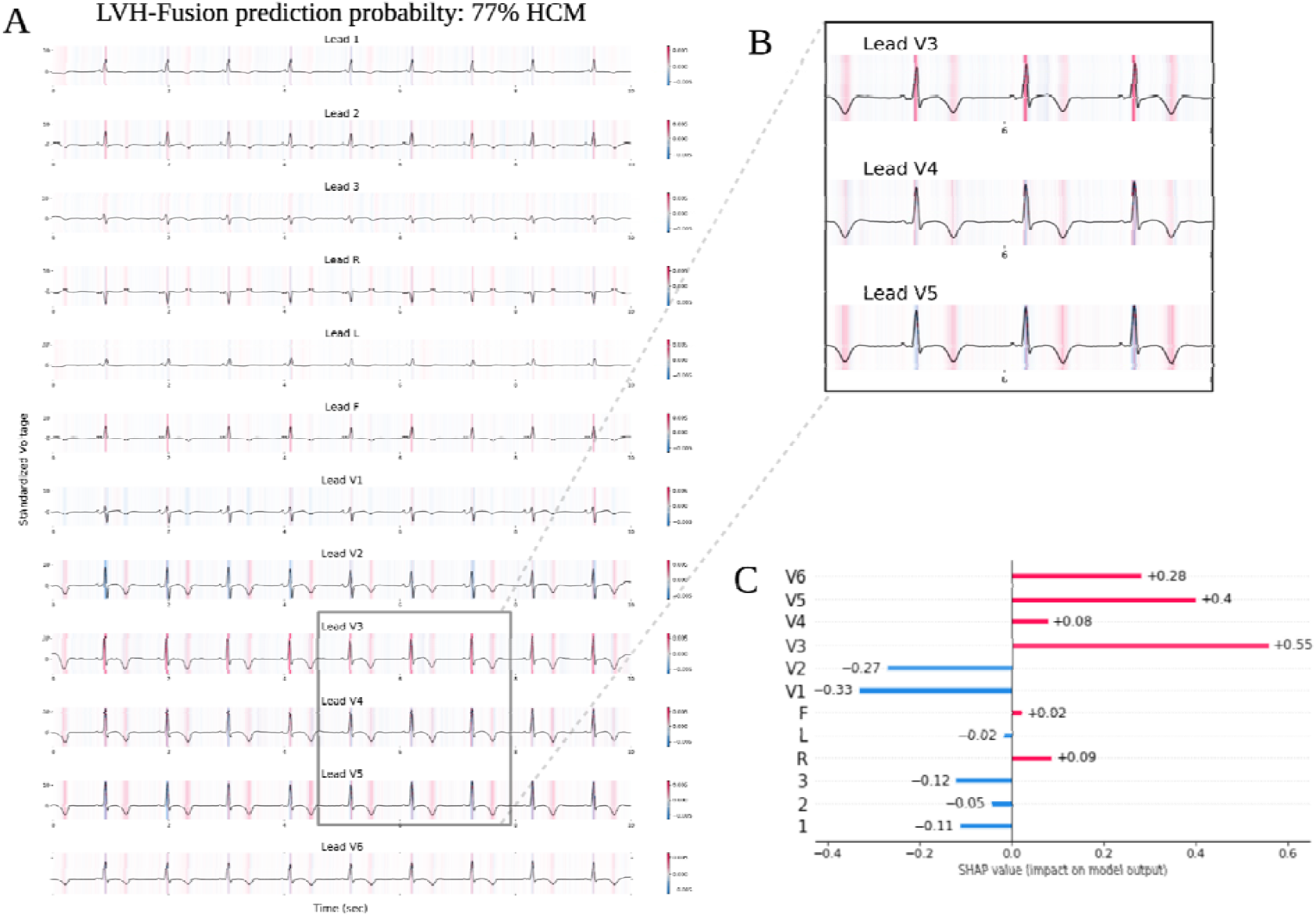
LVH-Fusion ECG interpretations. SHAP explanations of one true positive, HCM sample (A). Red areas indicate timesteps that hold a positive impact on prediction, while blue timesteps indicate a negative impact on prediction, no color is neutral. (B) Selected regions of ECG leads denote timesteps of high estimated importance, focusing on inverted T-waves and lead V3 R peaks. (C) Local explanations of the cumulative SHAP values on prediction output across leads. Lead V3 overall contains the highest values of SHAP values for this sample presented.

**Figure 4.**
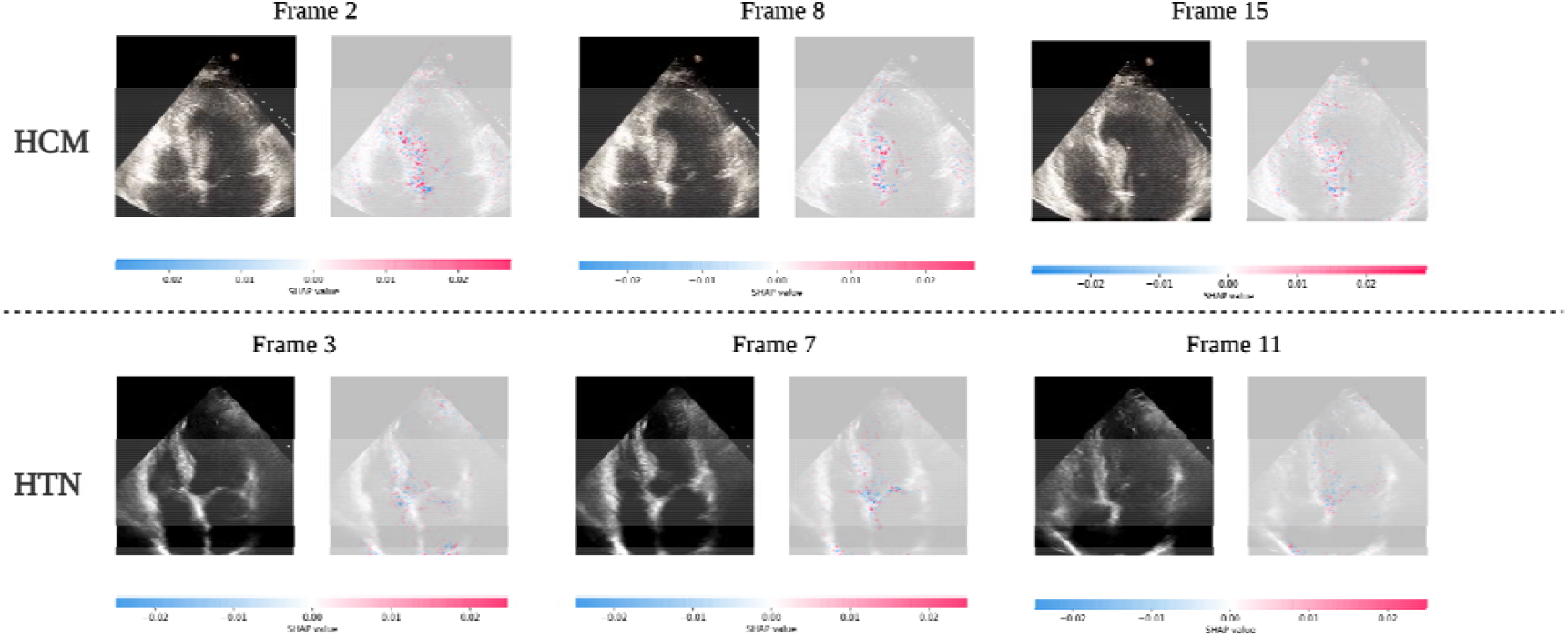
LVH-Fusion echocardiogram interpretations. SHAP explanations for two true positive samples, HCM (top row) and HTN (bottom row). Each class has 3 frames selected with SHAP values overlaid. Red areas indicate pixels that hold a positive impact on prediction, while blue pixels indicate a negative impact on prediction, no color is neutral. We observe red areas of importance converging on the asymmetric septal wall in the HCM example.

To further examine if the regions of importance identified in distinct samples are globally similar across all predictions, a summation or averaging across all local instances was performed. This approach provides a highly compressed, global insight into the model’s behavior. We considered per lead contributions to predictions in ECGs and left ventricular segmentation in echocardiogram videos. Global summary results for ECG corroborates our results from the ablation studies, lead V3 and aVR holds valuable information for model’s prediction estimates, Supplement Figure S4.

### Comparison against physician interpretation

We had two expert readers review ECG tracings and echocardiogram videos and asked them to make a diagnosis of HTN or HCM. We selected 45 samples (40 HTN and 5 HCM) from the test set to compare LVH-fusion. The LVH-fusion model outperformed these expert cardiologists (one of whom has 20 years of experience in diagnosing HCM). LVH-fusion correctly classified 3 out of the 5 ECG and echocardiogram HCM samples. Variability between cardiologists varied greatly, with one cardiologist matching LVH-fusion sensitivity estimates but with a reduction in specificity, while cardiologist two failed to correctly classify any of the HCM ECG samples provided.

## Discussion

In this study, we report the first multimodal (ECG and echocardiogram based) deep learning model in clinical cardiology and use it to predict the etiology of left ventricular hypertrophy. Combining complementary knowledge from multiple modalities can improve diagnostic performance in clinical practice. The trained model demonstrates high discriminatory ability in distinguishing hypertrophic cardiomyopathy from hypertension with an AUC of 0.91, AUPRC of 0.78. Furthermore, ablation studies provided independent support from unsupervised analysis for clinicians’ focus on ECG lateral repolarization and echocardiographic proximal septal hypertrophy for the diagnosis of HCM. Combining complementary information from multiple modalities is intuitively appealing for improving the performance of learning-based approaches. Our results can be directly applied in general medical and cardiology clinics where exposure to rare conditions such as HCM limits confidence in human diagnostic prediction alone.

Deep learning models specifically focused on single modalities in cardiology have shown impressive results for arrhythmia detection, age, and other clinical actionable insights ^8,10,16^. Previously Ko et al., focused on using convolutional neural networks (CNN) for ECG interpretation with respect to HCM ^22^. They showed high discriminatory power in classifying HCM against a background population of left ventricular hypertrophy by ECG alone. However, approximately 28-30% of HCM cases had concurrent hypertension, inhibiting a direct comparison of possible distinction between HCM and hypertension. To date, deep learning research addressing non-pulmonary hypertension detection using electrocardiogram or echocardiogram was unknown. One previous approach successfully used both ECG and echocardiogram data individually with a stepwise approach to diagnosis of cardiac amyloidosis^15^, whereas here we focus on fusion method applications of multi-modal deep learning of electrocardiograms and echocardiograms together.

Medical decision making is complex, often relying on a combination of physician’s judgment, experience, diagnostic and screening test results, and longitudinal follow-up. In the case of a patient presenting with anything other than severe, grossly asymmetric LVH, suspicion for HCM would be higher for patients who do not obviously have hypertension. However, occult hypertension is common and challenging to rule out and with mild “gray zone” hypertrophy, it is not uncommon to make this assumption. Similarly, for patients who present with LVH and manifest hypertension, the question is always “is hypertension alone enough to explain this degree of LVH?” Given the implications of missing a diagnosis of HCM—a mendelian disease associated with heart failure and sudden death—most generalists do not feel confident ignoring the possibility of HCM. In these cases, aggressively treating hypertension and re-reviewing the patient can help but challenges in follow up, adherence, and effectiveness of therapy make the window of equipoise long. These are the clinical scenarios into which LVH-fusion will have the most benefit. Yet, this is merely the first application of the approach. A similar approach to the identification of other causes of LVH such as Fabry disease or cardiac amyloidosis can be applied using similar “gold standard” diagnostic labels to those we use here. The future of deep learning in medicine is a move beyond reproducing human derived label features to capitalizing on unsupervised machine learned features vs a gold standard diagnostic or prognostic label. This will allow machine augmentation of the human led diagnostic journey.

In summary, we develop a deep learning model incorporating ECG and echocardiogram time series data and apply it to help identify hypertrophic cardiomyopathy patients from within the much larger group of patients presenting with LVH due to hypertension or unknown causes. We present various well known fusion methods of combining data streams from multiple modalities and compare these comprehensively to single-modal models. Further studies should explore the real-world application of physician augmentation approaches like LVH-fusion in medical practice.

## Methods

### Data acquisition and study population

Hypertrophic cardiomyopathy patients were selected for this study from the Hypertrophic Cardiomyopathy clinics at the Stanford Center for Inherited Cardiovascular Disease. Hypertension patients were selected from individuals that were found to be persistently hypertensive (SBP >150) with at least 5 consecutive systolic blood pressure readings over 150. Exclusion criteria included any ECG clinical annotations of ventricular-pacing or left bundle branch block. In addition, we excluded any data from both electrocardiograms and echocardiograms datasets if the date acquired was after a documented myectomy procedure.

We retrieved 15,761 electrocardiograms (ECGs) and 3,234 transthoracic echocardiograms from 2,728 unique individuals at Stanford Health Care, Table 1. Standard 12 lead ECGs were divided into training, validation, and test partitions based on a unique patient identification number to ensure that no patient overlap existed across data partitions. Echocardiogram videos from Stanford Medicine were curated for apical 4-chamber view videos.

### Data Processing and selection

Electrocardiogram signals were filtered to remove any baseline wander and powerline interference. Normalization of 12 lead ECGs was performed by lead over a random subset of the study sample population, using mean and standard deviation. Echocardiogram videos were processed in an identical method as Oyuang et al^13^. Given multiple electrocardiograms and echocardiograms per individual present within our dataset, we examined the effects of different data selection methods on model training and performance metrics. We selected three different data selection methods: 1) first clinical presentation for all data partitions, 2) all clinical presentations in the training partition with only first clinical presentation selected for the validation and test partitions, and 3) all clinical presentations for all partitions. Extended details of each selection method can be found in Supplemental Table 1.

### Overview of model training framework

Training for the single-modal and multimodal neural network models were executed independently.

Models were trained using a two-stage grid search approach to find the optimal hyperparameters. In the initial hyperparameter search, evaluation metrics from the validation set can be found in the Supplementary Tables S5, S6. The hyperparameters that yielded the best performing models were selected for additional training and hyperparameter search considering various loss functions, loss weighting for minority class and minority class oversampling. Final models were selected from the lowest validation loss.

### Single-modal model training

For electrocardiogram single-modal model training, the following hyperparameters included: model architecture: {VGG11, VGG13, VGG16, VGG19, densenet169, densenet121, densenet201, densenet161 resnet18, resnet34, resnet50, resnet101, resnet152, resnext50_32×4d, resnext101_32×8d, wide_resnet50_2 wide_resnet101_2}; batch size: {32, 64, 75}; Optimizer: {SGD, adam}, and Hz: {500, 250}. The first hyperparameter search involved training all combinations of hyperparameters above for 100 epochs and saving results from the epoch with the lowest loss. Furthermore, we explored a second hyperparameter search which explored class weighted loss functions, oversampling minority class samples and setting final bias term to the expected class ratios from top performing models from the initial hyperparameters search. We examined expanding training to 150 epochs and considering both loss and auPRC results for selection of the final model. The selected hyperparameters that resulted in best performance on the validation set were the following: resnet 34 model, oversampling minority class, adam optimizer, batch size of 64, and sampling rate of 500.

For echocardiogram unimodal model training, the following hyperparameters included: Model architecture: {r2plus1d_18, mc3_18, r3d_18}, Number of frames: {96, 64, 32, 16, 8, 4, 1}; Period: {2, 4}; Pretrained weights: {True, False}. The first hyperparameter search involved training all combinations of hyperparameters above for 100 epochs and saving results from the epoch with the lowest loss. Furthermore, we explored a second hyperparameter search which explored class weighted loss functions, oversampling minority class samples and setting final bias term to the expected class ratios from top performing models from the initial hyperparameters search. We examined expanding training to 300 epochs and considering both loss and auPRC results for selection of the final model. The selected hyperparameters that resulted in best performance on the validation set were the following: r2plus1d_18 model, pretrained weights, weighted minority class, adam optimizer, batch size of 20, and frames 16 with sampling period of 4.

### Multimodal model training

For multimodal training models, the electrocardiogram and echocardiogram data were paired according to unique patient identifiers. Data selection for the earliest clinical encounter was selected for all training, validation and test set partitions; this resulted in a total of 1,414 training, 176 validation, and 168 internal test samples. The detailed characteristics of the dataset can be found in Table 1. We hypothesized that using the learned weights from the single-modal models would benefit training so we explored both pre-trained late fusion and random late fusion models. All multimodal models were trained to 300 epochs and we considered both loss and auPRC results for selection of the final multimodal model. We implemented LVH-Fusion using PyTorch on the Stanford University Research cluster, Sherlock. The selected hyperparameters that resulted in best performance on the validation set were the following: r2plus1d_18 model + resnet 34, pretrained weights, weighted minority class, adam optimizer, batch size of 10, and frames 16 with sampling period of 4.

### Comparison to feature based models

Standard reported features from Tracemaster electrocardiogram machines were extracted for each ECG considered in this study. We used these features for input into a XGboost model to determine if a feature-based method would exceed the performance metrics of the unimodal neural network models. The list of ECG features used were modeled from Kwon et al. 2020 ^10^.

### Comparison with normal samples

In order to explore how our neural networks, perform on non-left ventricular hypertrophy individuals, we sampled electrocardiograms with clinical annotations of sinus rhythm and echocardiograms with a normal ejection fraction greater than 45. We took the best performing single-modal model and retrained them to include an additional non-LVH class; details of sample size and performance metrics can be found in Supplementary Table 5 and Supplementary Table 6, respectively.

### Ablation experiments

To further understand how the neural networks make their predictions, we explored various ablation studies.

We retrained the single-modal echo model with data ablated in the following ways:

1. a single randomly selected frame of each echo, repeated for the length of the original video to compare to the best performing unimodal model.
2. The end diastolic frame from each echo, repeated for the length of the original video to fairly compare to the best performing unimodal model. The end diastolic frame was identified by a trained sonographer from EchoNet-dynamic^13^.
3. Using the estimated left ventricular segmentation from EchoNet-dynamic^13^, we set all pixels to zero except a segmented box around the left ventricle.

For electrocardiogram we retrained the single-modal models for the following experiments:

1. Using 8 of the 12 leads, to compare to the best performing unimodal model.
2. Masking out each lead independently to compare to the best performing single-modal model and understand impacts each lead holds on performance.

Echocardiogram models were trained to 300 epochs and electrocardiogram models were trained for 150 epochs.

### SHAP Interpretation experiments

SHAP GradientExplainer^19^ uses an extension of integrated gradient values and SHAP values, which aims to attribute an importance value to each input feature by integrating the gradients of all interpolations between a foreground sample (test samples) and a provided background samples (training data). The importance scores sum up to approximately the difference between the expected value of all background samples and the individual prediction estimate of interest. We applied this method to both ECG and echocardiogram models; 1500 samples were used to build the background distribution for the ECG model and 80 samples were used to build the background distribution for the echocardiogram model. In both cases, the full test set was used as foreground samples.

## Data Availability

All the code for LVH-Fusion will be available at https://github.com/AshleyLab/lvh-fusion/ after publication. The data that support the findings of this study are available on request from the corresponding author upon approval of data sharing committees of the respective institutions.

## SUPPLEMENTAL TABLES AND FIGURES

**Supplemental Table S1.**

| Data Selection | Label | Number of unique Echo patients | Number of Echos | Number of unique ECG patients | Number of ECGs |
| --- | --- | --- | --- | --- | --- |
| First Encounters, train, val, and test | HCM | 314 | 314 | 811 | 811 |
|  | HTN | 1,836 | 1,836 | 1,917 | 1,917 |
| First Encounter, val and test | HCM | 314 | 654 | 811 | 4,430 |
|  | HTN | 1,836 | 2,343 | 1,917 | 8,730 |
| All Encounters train, val, and test | HCM | 324 | 763 | 917 | 6,027 |
|  | HTN | 1,848 | 2,516 | 1,977 | 13,045 |

**Supplemental Table S2.**

| Model | Loss | auPRC | Optim | Batch | Data Selection |
| --- | --- | --- | --- | --- | --- |
| VGG11 | 0.502 | 0.591 | adam | 75 | First Encounters, all |
| VGG13 | 0.448 | 0.672 | adam | 64 | First Encounters, all |
| VGG16 | 0.493 | 0.559 | adam | 64 | First Encounters, all |
| VGG19 | 0.501 | 0.559 | adam | 64 | First Encounters, all |
| densenet121 | 0.395 | 0.738 | adam | 64 | First Encounters, all |
| densenet161 | 0.449 | 0.639 | adam | 64 | First Encounters, all |
| densenet169 | 0.41 | 0.722 | adam | 75 | First Encounters, val and test only |
| densenet201 | 0.433 | 0.724 | adam | 64 | First Encounters, all |
| resnet101 | 0.427 | 0.702 | adam | 75 | First Encounters, all |
| resnet152 | 0.434 | 0.715 | adam | 64 | First Encounters, all |
| resnet18 | 0.405 | 0.727 | adam | 64 | First Encounters, all |
| resnet34 | 0.383 | 0.781 | adam | 64 | First Encounters, all |
| resnet50 | 0.429 | 0.73 | adam | 75 | First Encounters, all |
| resnext101_32x8d | 0.405 | 0.741 | adam | 75 | First Encounters, all |
| resnext50_32x4d | 0.405 | 0.734 | adam | 64 | First Encounters, all |
| wide_resnet101_2 | 0.426 | 0.69 | adam | 75 | First Encounters, all |
| wide_resnet50_2 | 0.416 | 0.692 | adam | 64 | First Encounters, all |

**Supplemental Table S3.**

| Model | loss | auPRC | optim | frames | period | File selection |
| --- | --- | --- | --- | --- | --- | --- |
| mc3 | 0.134 | 0.893 | adam | 16 | 2 | First Encounters, all |
| r2plus1d | 0.171 | 0.851 | adam | 16 | 4 | First Encounters, all |
| r3d | 0.152 | 0.879 | adam | 16 | 4 | First Encounters, all |

**Supplemental Table S4.**

|  | auROC | auPRC | F1score | Sensitivity | Specificity | PPV | NPV | FPR | FNR | FDR |
| --- | --- | --- | --- | --- | --- | --- | --- | --- | --- | --- |
| Reduced features (11) | 0.71 | 0.59 | 0.5 | 0.43 | 0.89 | 0.6 | 0.8 | 0.11 | 0.57 | 0.4 |
| Large features (468) | 0.84 | 0.73 | 0.59 | 0.47 | 0.95 | 0.78 | 0.82 | 0.05 | 0.53 | 0.22 |

**Supplemental Table S5.**

|  | f1-score | precision | recall | support |
| --- | --- | --- | --- | --- |
| HTN | 0.71 | 0.78 | 0.65 | 181 |
| HCM | 0.41 | 1.00 | 0.26 | 27 |
| NORMAL EF | 0.95 | 0.92 | 0.97 | 876 |
| macro avg | 0.69 | 0.90 | 0.63 | 1084 |
| weighted avg | 0.89 | 0.90 | 0.90 | 1084 |

**Supplemental Table S6.**

|  | f1-score | precision | recall | support |
| --- | --- | --- | --- | --- |
| HTN | 0.39 | 0.35 | 0.44 | 178 |
| HCM | 0.36 | 0.26 | 0.57 | 68 |
| SINUS | 0.91 | 0.95 | 0.88 | 1789 |
| macro avg | 0.56 | 0.52 | 0.63 | 2035 |
| weighted avg | 0.85 | 0.87 | 0.83 | 2035 |

**Supplemental Figure S1.**
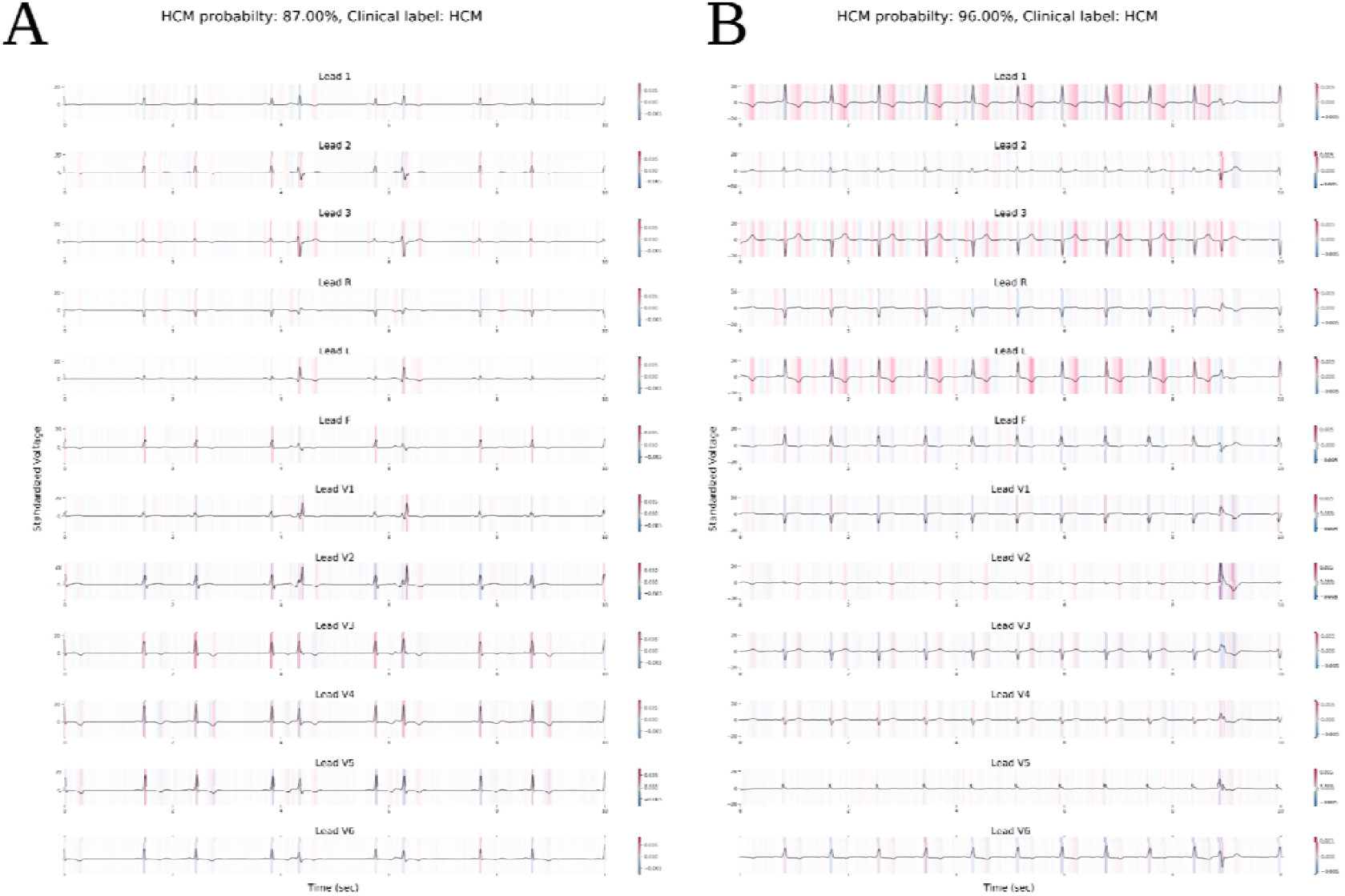
Additional examples of true positive HCM ECGs.

**Supplemental Figure S2.**
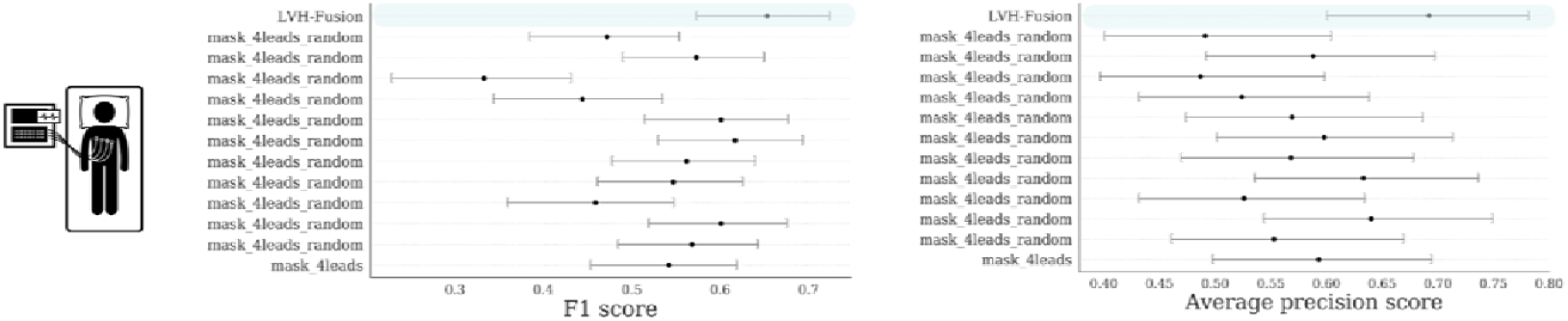
ECG ablation study of multiple lead masking.

**Supplemental Figure S3.**
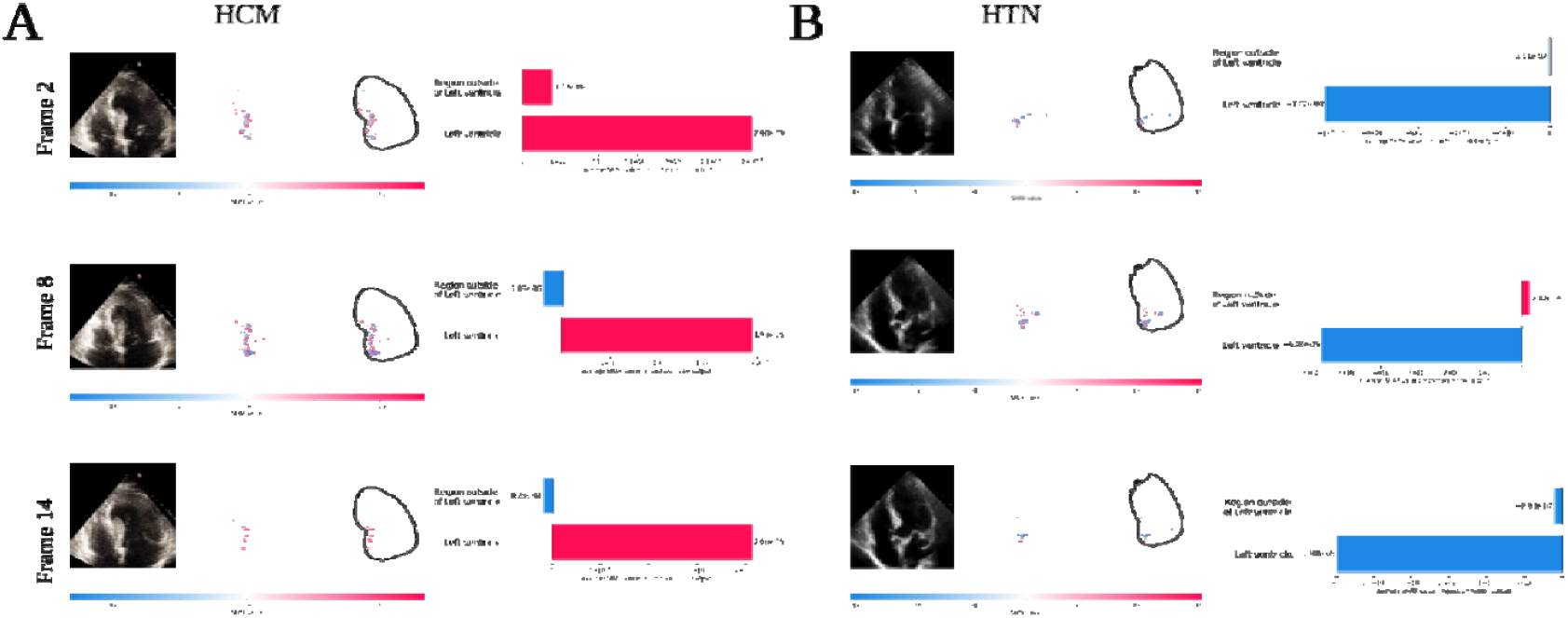
Echo SHAP local feature importance plot.

**Supplemental Figure S4.**
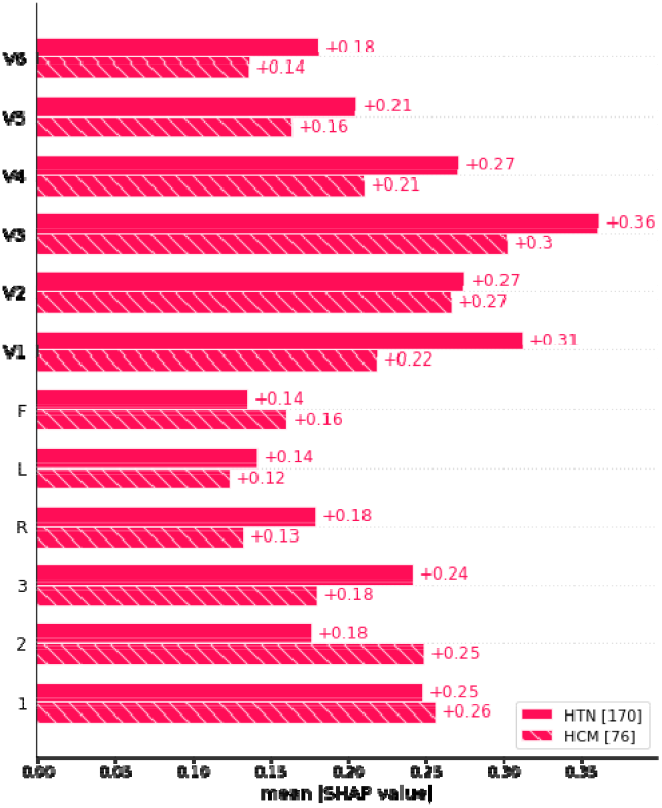
ECG SHAP global feature importance plot. The global importance of each lead is taken to be the mean absolute value summation for each lead over all the given samples. Hypertension (HTN) is in solid red, Hypertrophic Cardiomyopathy (HCM) is denoted by stripes. Lead V3 is ranks highest overall in global feature importance.

## References

1. Semsarian, C., Ingles, J., Maron, M. S. & Maron, B. J. New perspectives on the prevalence of hypertrophic cardiomyopathy. J. Am. Coll. Cardiol. 65, 1249–1254 (2015).

2. Ho, C. Y. et al. Genotype and Lifetime Burden of Disease in Hypertrophic Cardiomyopathy: Insights from the Sarcomeric Human Cardiomyopathy Registry (SHaRe). Circulation 138, 1387–1398 (2018).

3. Whelton, P. K. et al. 2017 ACC/AHA/AAPA/ABC/ACPM/AGS/APhA/ASH/ASPC/NMA/PCNA Guideline for the Prevention, Detection, Evaluation, and Management of High Blood Pressure in Adults: A Report of the American College of Cardiology/American Heart Association Task Force on Clinical Practice Guidelines. Hypertension 71, e13–e115 (2018).

4. Magnusson, P., Palm, A., Branden, E. & Mörner, S. Misclassification of hypertrophic cardiomyopathy: validation of diagnostic codes. Clin. Epidemiol. 9, 403–410 (2017).

5. Esteva, A. et al. Deep learning-enabled medical computer vision. NPJ Digit Med 4, 5 (2021).

6. Pennacchini, E., Musumeci, M. B., Fierro, S., Francia, P. & Autore, C. Distinguishing hypertension from hypertrophic cardiomyopathy as a cause of left ventricular hypertrophy. J. Clin. Hypertens. 17, 239–241 (2015).

7. Doi, Y. L. et al. Echocardiographic differentiation of hypertensive heart disease and hypertrophic cardiomyopathy. Br. Heart J. 44, 395–400 (1980).

8. Attia, Z. I. et al. Age and Sex Estimation Using Artificial Intelligence From Standard 12-Lead ECGs. Circ. Arrhythm. Electrophysiol. 12, e007284 (2019).

9. Hannun, A. Y. et al. Cardiologist-level arrhythmia detection and classification in ambulatory electrocardiograms using a deep neural network. Nat. Med. 25, 65–69 (2019).

10. Kwon, J.-M. et al. A deep learning algorithm to detect anaemia with ECGs: a retrospective, multicentre study. Lancet Digit Health 2, e358–e367 (2020).

11. Yao, X. et al. Artificial intelligence–enabled electrocardiograms for identification of patients with low ejection fraction: a pragmatic, randomized clinical trial. Nature Medicine (2021) doi:10.1038/s41591-021-01335-4.

12. Madani, A., Ong, J. R., Tibrewal, A. & Mofrad, M. R. K. Deep echocardiography: data-efficient supervised and semi-supervised deep learning towards automated diagnosis of cardiac disease. NPJ Digit Med 1, 59 (2018).

13. Ouyang, D. et al. Video-based AI for beat-to-beat assessment of cardiac function. Nature 580, 252–256 (2020).

14. Huang, S.-C., Pareek, A., Zamanian, R., Banerjee, I. & Lungren, M. P. Multimodal fusion with deep neural networks for leveraging CT imaging and electronic health record: a case-study in pulmonary embolism detection. Sci. Rep. 10, 22147 (2020).

15. Goto, S. et al. Artificial intelligence-enabled fully automated detection of cardiac amyloidosis using electrocardiograms and echocardiograms. Nat. Commun. 12, 2726 (2021).

16. Attia, Z. I. et al. An artificial intelligence-enabled ECG algorithm for the identification of patients with atrial fibrillation during sinus rhythm: a retrospective analysis of outcome prediction. Lancet 394, 861–867 (2019).

17. Sagi, O. & Rokach, L. Ensemble learning: A survey. Wiley Interdiscip. Rev. Data Min. Knowl. Discov. 8, e1249 (2018).

18. Hughes, J. W. et al. Deep learning prediction of biomarkers from echocardiogram videos. bioRxiv (2021) doi:10.1101/2021.02.03.21251080.

19. Sundararajan, M., Taly, A. & Yan, Q. Axiomatic attribution for deep networks. arXiv [cs.LG] (2017).

20. Lewington, S. et al. Age-specific relevance of usual blood pressure to vascular mortality: a meta-analysis of individual data for one million adults in 61 prospective studies. Lancet 360, 1903–1913 (2002).

21. Parato, V. M. et al. Echocardiographic diagnosis of the different phenotypes of hypertrophic cardiomyopathy. Cardiovasc. Ultrasound 14, 30 (2016).

22. Ko, W.-Y. et al. Detection of Hypertrophic Cardiomyopathy Using a Convolutional Neural Network-Enabled Electrocardiogram. J. Am. Coll. Cardiol. 75, 722–733 (2020).

